# Low-coverage genome sequencing for the detection of clinically relevant copy-number and mtDNA variants

**DOI:** 10.1101/2022.09.20.22280155

**Authors:** Sander Pajusalu, Mikk Tooming, Kaisa Teele Oja, Ustina Šamarina, Tiina Kahre, Katrin Õunap

## Abstract

**Background:** Compared to exome sequencing, genome sequencing is widely appreciated for its superior ability to detect a wide range of genetic variations including copy-number variants (CNVs) and mitochondrial (mtDNA) variants. We assessed whether low-coverage genome sequencing, a considerably cheaper approach, would detect clinically relevant CNVs and mtDNA variants and would thus be a cost-efficient supplement to exome sequencing in rare disease diagnostics.

**Methods:** To assess the level of sequencing depth needed for variant detection, first, 30x mean coverage genome sequencing data were subsampled to 0.5x, 1x, 2x, and 4x coverage files *in silico* followed by CNV and mtDNA detection. Based on the analysis, 2x short-read sequencing was selected to be performed in 16 patients with putatively pathogenic CNVs or mtDNA variants to assess the empirical sensitivity.

**Results:** For CNV calling, 2x coverage was sufficient to detect all heterozygous CNVs greater than 10kb in size from *in silico* subsampled data. In experimental data, the results were similar, although a 16kb heterozygous deletion was once not detected. Regarding mtDNA variants, 2x coverage sufficed for variant confident variant calling and heteroplasmy assessment for all samples.

**Conclusions:** Low-coverage genome sequencing may be used to complement exome sequencing for simultaneous mtDNA variant and CNV detection.

## Introduction

Since the first human exome sequencing study focused on finding a genetic cause for rare human diseases (Ng et al., 2009), novel sequencing technologies facilitating genome-wide simultaneous variant detections have revolutionized research and diagnostics of genetic disorders (Wright et al., 2018).

Currently, many diagnostic labs are performing exome sequencing and chromosomal microarray (CMA) in parallel to discover most of the clinically interpretable findings, as exome sequencing can detect single nucleotide variants and short deletions and insertions, while CMA is developed for detecting copy-number variants (CNVs), which are causative factors for ∼10% of rare genetic disorders (Žilina et al., 2014). In case of suspicion of mitochondrial disorders, also mtDNA sequencing is requested.

High-depth genome sequencing can outperform CMAs in sensitivity for CNV detection, as it is not limited to the size resolution and can effectively detect both copy-number variants and balanced structural variants (e.g., translocations and inversions) (Collins et al., 2020). However, high computational and reagent costs challenge the usage in clinical settings. Generally, high-depth genome sequencing at standard 30x coverage is 3-5 times more expensive than exome sequencing, while CMA is cheaper than exome. Low coverage genome sequencing has been proposed as an alternative for CMA, as the lower coverage will reduce the costs proportionally while analytical sensitivity still outperforms CMA, especially for smaller deletions and duplications even at only 1x coverage (Dong et al., 2016; Zhou et al., 2018). Low-coverage genome sequencing has also been tested in prenatal settings (Wang et al., 2020). Several read-depth-based computational tools have been used for CNV detection from low-coverage genome sequencing data. Control-FREEC (Boeva et al., 2012) has shown the best performance with optimal computational resource usage (Smolander et al., 2021).

Another way to increase diagnostic yield of next-generation sequencing is to simultaneously detect mitochondrial DNA (mtDNA) variants, as exome and genome sequencing also cover mtDNA as a byproduct (Duan et al., 2018, 2019). An average PCR-free clinical genome sequencing has a mean read depth of around 30x, which results in above 2000x mtDNA coverage due to a large copy number of mtDNA in cells compared to autosomes (Laricchia et al., 2022). By reducing genome depth, sequencing costs decrease proportionally, but mtDNA coverage, although lower, could still be sufficient for variant detection. A previous study demonstrated that average autosome coverage of 1.6x resulted in average mtDNA coverage of 124x on genome sequencing (Rustagi et al., 2017). The sequencing depth of 100x or more is sufficient for detecting variants with heteroplasmy (proportion of mtDNA molecules having the non-reference allele) levels over 10% covering all clinically relevant variants if DNA from disease-relevant tissue is sequenced.

This study aims to assess whether low-coverage genome sequencing could be a reasonably cost-efficient solution for detecting CNVs and mtDNA variants and thus supplementing exome sequencing.

## Methods

First, to assess the level of sequencing depth needed for variant detection, 30x mean coverage genome sequencing data from selected samples were subsampled to 0.5x, 1x, 2x, and 4x coverage files *in silico*. CNVs were detected using Control-FREEC (Boeva et al., 2012) and annotated with AnnotSV (Geoffroy et al., 2018). The mtDNA variants were detected using the GATK4 mitochondrial pipeline (Laricchia et al., 2022) and annotated with HmtNote (Preste et al., 2019). The GATK4 mitochondrial pipeline also outputs theoretical sensitivity assessment for different heteroplasmy levels, which was used for selecting genome sequencing depth for the second part of the study. Five disease-causing deletions (sized 3.4kb, 4.6kb, 7.2kb, 16kb, and 90kb) and one possibly pathogenic heteroplasmic (heteroplasmy level 14.7%) mtDNA variant were used to assess sensitivity for clinically relevant variants.

Second, 16 samples from ten different families with known variants were selected for 2x genome sequencing. The chosen samples carried the following variants: eight (five unique) deletions, one duplication, three (one unique) inversions, and four (three unique) mtDNA variants (Tables 1 and 2). The sequencing run was carried out on Illumina NextSeq 500 in a single run using high output sequencing kit with 2×150bp paired-end reads. Fastq files were mapped to the hg38 reference genome using BWA MEM algorithm version 0.7.17-r1188 (Li & Durbin, 2009), duplicates were marked, and base quality scores recalibrated using GATK version 4.1.4.0 (van der Auwera et al., 2020). For quality control, genome sequencing metrics, including sequencing depth, were assessed with the Picards CollectWgsMetrics tool (http://broadinstitute.github.io/picard/). A read-depth assessment-based Control-FREEC software (Boeva et al., 2012) with 1kb and 10kb non-overlapping calling windows and Manta, combining paired and split-read evidence (Chen et al., 2016) was used to call structural variants from bam files. GATK4 mitochondrial pipeline (Laricchia et al., 2022) was used to detect mtDNA variants and assess heteroplasmy. Heteroplasmy was calculated as a ratio of alternate variant reads to the total sequencing depth for the same genome locus. Again, AnnotSV (Geoffroy et al., 2018) and HmtNote (Preste et al., 2019) were used to annotate structural and mtDNA variants, respectively.

**Table 1.** Cohort of patients with known structural variants selected for 2x genome sequencing (GS) and the results with 2 different variant callers. Control-FREEC software was used in two modes using either one kilobase or ten kilobase windows. Fam – family, DEL – deletion, DUP – duplication, INV – inversion.

| Fam | Relation | Known variant from 30x GS | Mean depth | Control-FREEC 10kb window | Control-FREEC 1kb window | Manta |
| --- | --- | --- | --- | --- | --- | --- |
| 1 | Proband | HOM 16kb DEL in *WBP4* | 2.12 | Detected | Detected | Not detected |
| 1 | Mother | HET 16kb DEL in *WBP4* | 2.05 | Not detected | Not detected | Not detected |
| 1 | Father | HET 16kb DEL in *WBP4* | 1.82 | Detected | Not detected | Not detected |
| 2 | Proband | HET 3.4kb DEL in *CTCF* | 1.94 | Not detected | Not detected | Not detected |
| 3 | Proband | HET 7.19kb DEL in *TRAPPC9* | 1.86 | Not detected | Not detected | Detected |
| 3 | Father | HET 7.19kb DEL in *TRAPPC9* | 1.86 | Not detected | Not detected | Not detected |
| 4 | Proband | HET 90.1kb DEL in *RBFOX1* | 1.82 | Detected | Detected | Not detected |
| 5 | Proband | HET 4.63kb DEL in *NIPBL* | 1.98 | Not detected | Not detected | Not detected |
| 6 | Proband | HET 14.2kb DUP in *SQOR* | 1.95 | Not detected | Detected | Detected |
| 7 | Proband | HOM 9Mb INV on chr 9 | 1.62 | Not applicable | Not applicable | Detected |
| 7 | Mother | HET 9Mb INV on chr 9 | 1.78 | Not applicable | Not applicable | Not detected |
| 7 | Father | HET 9Mb INV on chr 9 | 1.74 | Not applicable | Not applicable | Detected |

**Table 2.** Cohort of patients with mitochondrial DNA variants selected for 2x genome sequencing (GS) and the results for the known variants.

| Fam | Relation | Known mtDNA variant | Mean genome depth | Mean mtDNA depth | Heteroplasmy from 2x GS |
| --- | --- | --- | --- | --- | --- |
| 8 | Proband | m.9176T>C<br>95% heteroplasmy | 2.16 | 166 | 93.8% (183/195) |
| 9 | Proband | m.3243A>G<br>9.5% heteroplasmy | 1.79 | 202 | 7% (15/213) |
| 10 | Proband | m.15866A>G<br>14.7% heteroplasmy | 2.07 | 299 | 17% (60/352) |
| 10 | Mother | m.15866A>G<br>5.6% heteroplasmy | 2 | 225 | 4.5% (11/246) |

The scripts used for the analysis are available at https://github.com/SanderEST/lcwgs.

This study was approved by the Research Ethics Committee of the University of Tartu (approval date 11/18/2018 and number 287M-15, and 19/10/2020 327T-3). Informed consent was obtained from patients or their legal guardians.

## Results

First, in-silico subsampled data was assessed. For CNV calling, 2x coverage was sufficient to detect all heterozygous CNVs greater than 10kb. For smaller CNVs, even 4x coverage data did not suffice for CNV detection. An example of an estimated copy number using 10 kb windows on chromosome 13 around 16 kb deletion in both heterozygous and homozygous states is shown for different *in silico* subsampled depths in Figure 1. Regarding mtDNA variants, 2x coverage resulted in >99% theoretical sensitivity for heteroplasmy levels >10%. The possibly pathogenic heteroplasmic variant was detected with similar heteroplasmy levels in all depths (0.5x to 4x). We selected 2x as an aimed depth for the separate low-coverage genome sequencing experiment based on these results.

**Figure 1.**
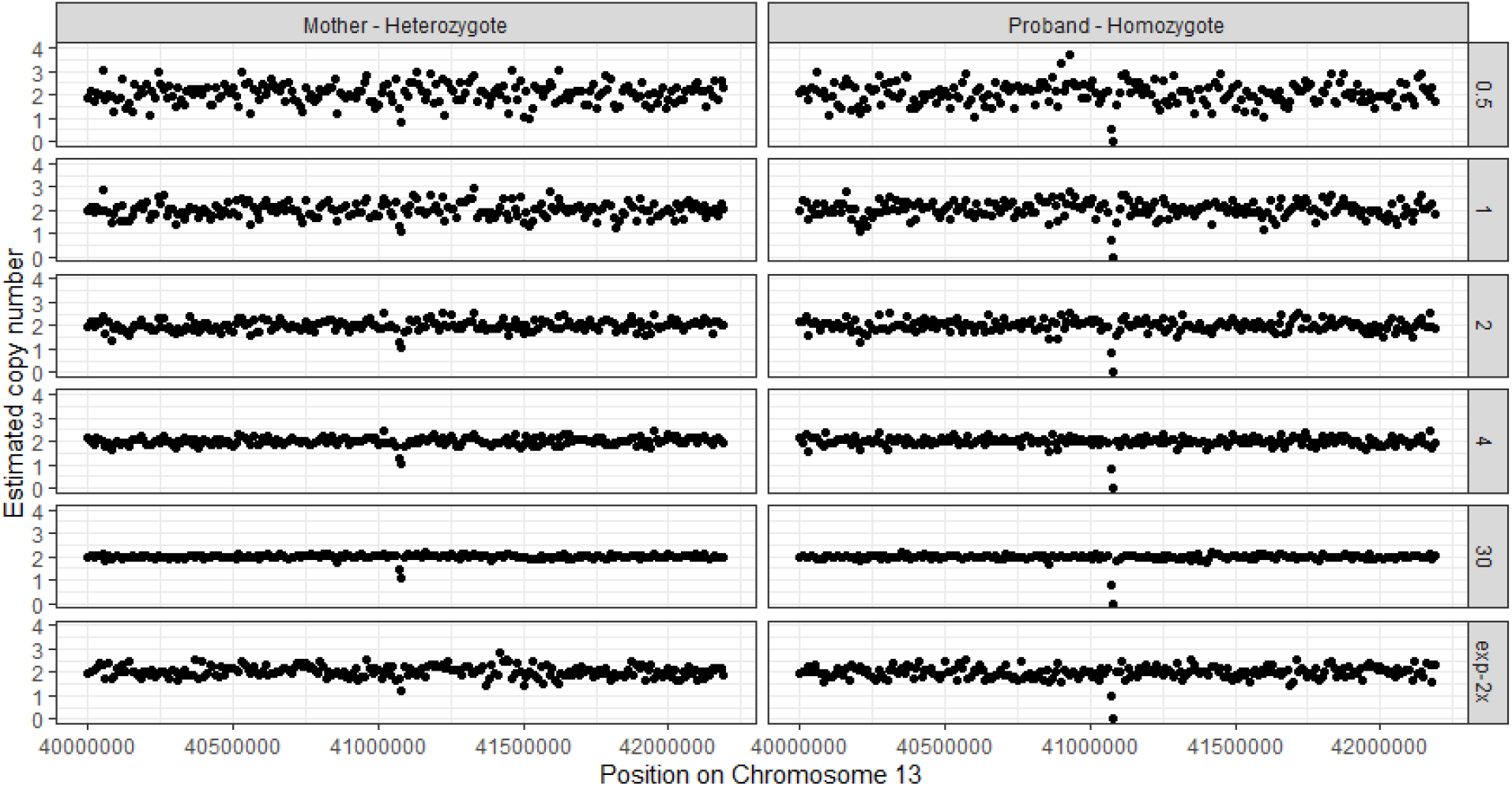
An example of an estimated copy number using 10 kb windows on chromosome 13 around 16 kb deletion in both heterozygous and homozygous states is shown for different *in silico* subsampled depths (0.5x, 1x, 2x, 4x) and the original 30x genome sequencing data as well as from the experimental 2x genome sequencing run (exp-2x).

The sequencing depth for 16 samples selected for the experiment ranged from 1.62 to 2.12, following the aimed sequencing depth of 2x. Regarding the assessed variants, FREEC confidently detected 90kb heterozygous deletion and 16kb homozygous deletion (Table 1, Figure 1). The detection was inconsistent for heterozygous 16kb deletion, and the software failed to detect smaller than 10kb deletions. Manta, using different algorithms, detected the variants with incomplete sensitivity which was not in direct concordance with the CNV size. For example, it was able to detect 7.2 kb deletion, and 14 kb duplication, which both were not detected by FREEC software, but was not able to detect even 90kb deletion and 16 kb homozygous deletion. We also assessed Manta’s ability to detect large inversion on chromosome 9, which was detected in the homozygous state, and in one of the two heterozygous carriers.

The known putatively pathogenic mtDNA variants were all detected from 2x genome sequencing. Moreover, the heteroplasmy levels were concordant with the 30x genome data (Table 2).

## Discussion

Although the field of rare disease diagnostics and research is shifting toward using high-depth genome sequencing as a first-tier test, the high cost for sequencing and computational demands make exome sequencing the most widely used test. Although possible, CNV detection from exome sequencing is challenged by the fragmented nature of the data (Pfundt et al., 2016). Thus, chromosomal microarrays are often used to supplement exome sequencing to detect clinically relevant CNVs. The detection limit of chromosomal microarrays depends on the array used, commonly ranging from 10kb to 100kb.

Also, mtDNA variants are often assessed separately. Although possible to detect from exome sequencing, the coverage is often poor and insufficient for heteroplasmy level assessment (Puusepp et al., 2018). Thus, a patient with a suspected genetic disorder but without a specific diagnostic hypothesis commonly receives three separate genetic tests, exome, mtDNA sequencing, and chromosomal microarray, making comprehensive testing expensive.

This study demonstrates that low-coverage genome sequencing can replace chromosomal microarray and mtDNA sequencing. However, some limitations have to be noted. For chromosomal microarrays, the resolution, i.e., the smallest size of a CNV that can be reliably detected, is provided by the manufacturer after sensitivity assessments. Similarly, low-coverage genome sequencing has its resolution, which may depend on the sequencing protocol and the bioinformatics pipeline. Each lab should assess the sensitivity and specificity of its protocol. Importantly, natural variation of sequencing depth should be considered as the depth for samples in the same run is never equal (Table 1).

Regarding mtDNA variant detection, the main limitation lies in the studied tissue. Exome sequencing is usually performed from the DNA extracted from the blood or saliva. In contrast, muscle or fibroblasts may be the preferred tissue for mtDNA variant detection due to differences in heteroplasmy levels between tissues. This should be noted, as it is tempting to use the already available DNA from exome sequencing for further studies. While the low heteroplasmy levels may not be detected from the low coverage genome sequencing, the sensitivity for heteroplasmy levels above 10% remains adequate. Thus this method is suitable for screening clinically relevant mtDNA variants.

As high-coverage genome sequencing is becoming cheaper, low-coverage genome sequencing may not be efficient in the future, where standard genome sequencing replaces exome sequencing, and other variant classes may be assessed from the same data. High coverage genome sequencing is probably more sensitive for other variant classes like repeat expansion variant detection (Ibañez et al., 2022). However, for the next few years, low coverage genome sequencing can serve as a cost-effective complementing analysis for exome sequencing, widening the scope of variant detection.

## Conclusions

Low-coverage genome sequencing may be used to complement exome sequencing for simultaneous mtDNA variant and structural variant detection. However, for smaller CNVs, higher coverage genome sequencing is needed for comprehensive variant detection.

## Data Availability

All data produced in the present study are available upon reasonable request to the authors.

## Acknowledgments

This study was supported by European Regional Development Fund and the program Mobilitas Pluss grant MOBTP175. KÕ, TK, KTO, and MT received support from the Estonian Research Council grants PRG471 and PUT355. The high coverage genome sequencing used for methods development was provided by the Broad Institute of MIT and Harvard Center for Mendelian Genomics (Broad CMG) and was funded by the National Human Genome Research Institute, the National Eye Institute, and the National Heart, Lung and Blood Institute grant UM1 HG008900 and in part by National Human Genome Research Institute grant R01 HG009141.

